# Convalescent plasma for preventing critical illness in COVID-19: A phase 2 trial and immune profile

**DOI:** 10.1101/2021.02.16.21251849

**Authors:** Jeffrey M. Sturek, Tania A. Thomas, James D. Gorham, Chelsea A. Sheppard, Allison E. Raymond, Kristen Petros De Guex, William B. Harrington, Andrew J. Barros, Gregory R. Madden, Yosra M. Alkabab, David Lu, Qin Liu, Melinda D. Poulter, Amy J. Mathers, Archana Thakur, Ewa M. Kubicka, Lawrence G. Lum, Scott K. Heysell

## Abstract

**Rationale:** The COVID-19 pandemic caused by the severe acute respiratory syndrome coronavirus 2 (SARS-CoV-2) is an unprecedented event requiring rapid adaptation to changing clinical circumstances. Convalescent immune plasma (CIP) is a promising treatment that can be mobilized rapidly in a pandemic setting.

**Objectives:** We tested whether administration of SARS-CoV-2 CIP at hospital admission could reduce the rate of ICU transfer or 28 day mortality.

**Methods:** In a single-arm phase II study, patients >18 years-old with respiratory symptoms documented with COVID-19 infection who were admitted to a non-ICU bed were administered two units of CIP within 72 hours of admission. Detection of respiratory tract SARS-CoV-2 by polymerase chain reaction and circulating anti-SARS-CoV-2 antibody titers were measured before and at time points after CIP transfusion.

**Measurements and Main Results:** Twenty-nine patients were transfused CIP and forty-eight contemporaneous controls were identified with comparable baseline characteristics. Levels of anti-SARS-CoV-2 IgG, IgM, and IgA anti-spike, anti-receptor-binding domain, and anti-nucleocapsid significantly increased from baseline to post-transfusion for all proteins tested. In patients transfused with CIP, the rate of ICU transfer was 13.8% compared to 27.1% for controls with a hazard ratio 0.506 (95% CI 0.165-1.554), and 28-day mortality was 6.9% compared to 10.4% for controls, hazard ratio 0.640 (95% CI 0.124-3.298).

**Conclusions:** Transfusion of high-titer CIP to patients early after admission with COVID-19 respiratory disease was associated with reduced ICU transfer and 28-day mortality but was not statistically significant. Follow up randomized trials may inform the use of CIP for COVID-19 or future coronavirus pandemics.

## INTRODUCTION

Passive antibody infusion was the first immunotherapy dating back to the 1890s for the treatment of infectious diseases before the development of antibiotics (1, 2). Experience from prior outbreaks with other coronaviruses, such as SARS-CoV-1 shows that such convalescent immune plasma (CIP) contains neutralizing antibodies to the relevant virus (3). The use of CIP or hyperimmune immunoglobulin-containing specific anti-viral antibodies has been safe and effective with other respiratory viral infections (4); and in the most recent Ebola outbreak, treatment with neutralizing antibodies improved survival when compared to pharmacologic inhibition (5).

Despite most studies demonstrating trends toward improvement in clinical endpoints for COVID-19 in patients treated with CIP, the rapid expansion of compassionate use of CIP for hospitalized patients in the US early in the COVID-19 pandemic made it challenging to design, coordinate, and obtain funding for randomized controlled trials (6). In March 2020, the US Food and Drug Administration (FDA) authorized CIP for compassionate use and in April, the FDA approved an expanded access program (EAP) for use of CIP for hospitalized patients at-risk of critical illness or those critically ill with COVID-19. CIP received emergency use authorization (EUA) from the FDA in August 2020. CIP remains one of the few therapies to have EUA for hospitalized patients along with the anti-viral remdesivir, with or without the selective inhibitor of Janus kinase 1 and 2, baricitinib (7). CIP has also been commonly considered as a bridge to specific monoclonal antibody therapies. While such therapies have demonstrated efficacy in prevention of medically attended visits among outpatients with COVID-19 (8, 9), a recent study of the monoclonal antibody bamlanivimab did not show sustained recovery in hospitalized patients, and the study was stopped early for futility (10). The failure of a monoclonal antibody in this population suggests that CIP containing polyclonal antibodies targeting multiple antigens may have a clinical advantage. Given the rapidity with which CIP can be mobilized, studies that confirm safety and efficacy can inform its role in future coronavirus pandemics (11).

The primary objectives of this phase II single-arm study were to determine if early administration of high titer SARS-CoV-2 CIP to adults hospitalized with respiratory symptoms from COVID-19 would be safe, and whether it would decrease transfer to the intensive care unit (ICU) and 28 day mortality. Secondary endpoints were to determine the kinetics of the development of specific antibody titers to SARS-CoV-2 viral proteins and determine correlation with specific antibody levels and the persistence of virus.

## METHODS

The Convalescent Immune Plasma -Treatment (CIP-T) trial is a single-arm, phase II study of CIP for the treatment of severe COVID-19 (hospitalized mild/moderately-ill, non-ICU, meeting World Health Organization (WHO) Ordinal Scale of 3 or 4) (12) patients compared to contemporaneous controls, with primary endpoints of 28 day mortality and ICU admission. This single-site study was performed at the University of Virginia Hospital, an academic medical center with a large referral base throughout the state. Pre-screened, high-titer convalescent plasma was provided by the New York Blood Center. Randomized placebo controls were deferred, considering limited evidence suggesting potential benefit in COVID-19 infection and the FDA EAP which was active at the time of study initiation.

An enrollment target of 29 patients was calculated to provide 80% power to detect a 50% reduction in the primary outcome using early historical targets of ICU admission from reports in China and elsewhere (13, 14). This study received approval from the University of Virginia Institutional Review Board (#200114), was conducted under the FDA IND BB-20867, and registered at clinicaltrials.gov, NCT04374565. Additional details with regard to convalescent plasma testing, laboratory measurements, clinical follow up, and statistical analysis can be found in the supplemental material.

### Study Population

Inclusion criteria were patients >18 years with respiratory symptoms attributable to COVID-19 (confirmed by positive oropharyngeal or nasopharyngeal SARS-CoV-2 PCR testing) within 72 hours of admission to an acute care (non-ICU) bed. People who had received chloroquine derivatives were eligible but were required to be taken off of the drug prior to enrollment. Participants could receive remdesivir and corticosteroids if not part of another anti-COVID-19 trial. Permissive inclusion criteria were set to achieve representative accrual of those with kidney or liver injury, or those with other comorbidities that may have excluded them from other COVID-19 therapeutic trials.

Exclusion criteria were mechanical ventilation or >6 liters per minute nasal cannula oxygen requirement, enrollment into other anti-COVID-19 trials available at our institution (which included the ACTT-1 and ACTT-2 trials), prior administration of tocilizumab (anti-IL-6 receptor) or siltuximab (anti-IL-6), or presence of a pre-existing condition, which, in the opinion of the site investigator, could place the individual at substantially increased risk of transfusion-related complications, such as the risk of volume overload with decompensated heart failure, or a history of prior transfusion reaction.

### Controls

To select contemporaneous controls that could have been feasibly enrolled and treated with CIP in this study, we screened adult patients with confirmed COVID-19 who were admitted for >48h during the study period (April 4 – August 16, 2020), and outside of an ICU setting for the first 12 hours. Controls were then reviewed by clinicians with expertise in Pulmonary & Critical Care and/or Infectious Diseases (JMS, SKH, TAT) who were provided History and Physical documentation, but were blinded to patient outcomes. Eligible controls were included in the final analysis if two clinician reviewers agreed the patient met study enrollment criteria including respiratory symptoms attributable to COVID-19 infection. Discordant assessments were adjudicated by a third reviewer.

## RESULTS

### Clinical Outcomes

Thirty-two participants were enrolled to reach the target of 29 that were ultimately transfused with CIP and included in the final analysis. One participant was discharged within a day of enrollment and prior to receipt of CIP, while two other participants signed consent but later declined CIP and were discharged alive. Among 149 potential controls screened, 48 met eligibility (Table 1). Participants that received CIP and controls were well-balanced, without statistically significant differences in demographics or co-morbidities. The mean age of participants and controls were 57.6 years (SE 2.5) and 64.4 years (SE 2.6), respectively (p=0.13). Fourteen (48%) participants were female compared to 30 (63%) controls (p=0.24). People of Black race or Hispanic ethnicity comprised the majority in both groups, including 19 (66%) participants and 36 (75%) controls, reflecting regional COVID-19 demographics. The most common comorbidity was hypertension, present in 17 (59%) participants and 31 (65%) controls (p=0.63). Participants that received CIP had higher admission levels of D-dimer and ferritin but these differences did not reach statistical significance.

**Table 1.**
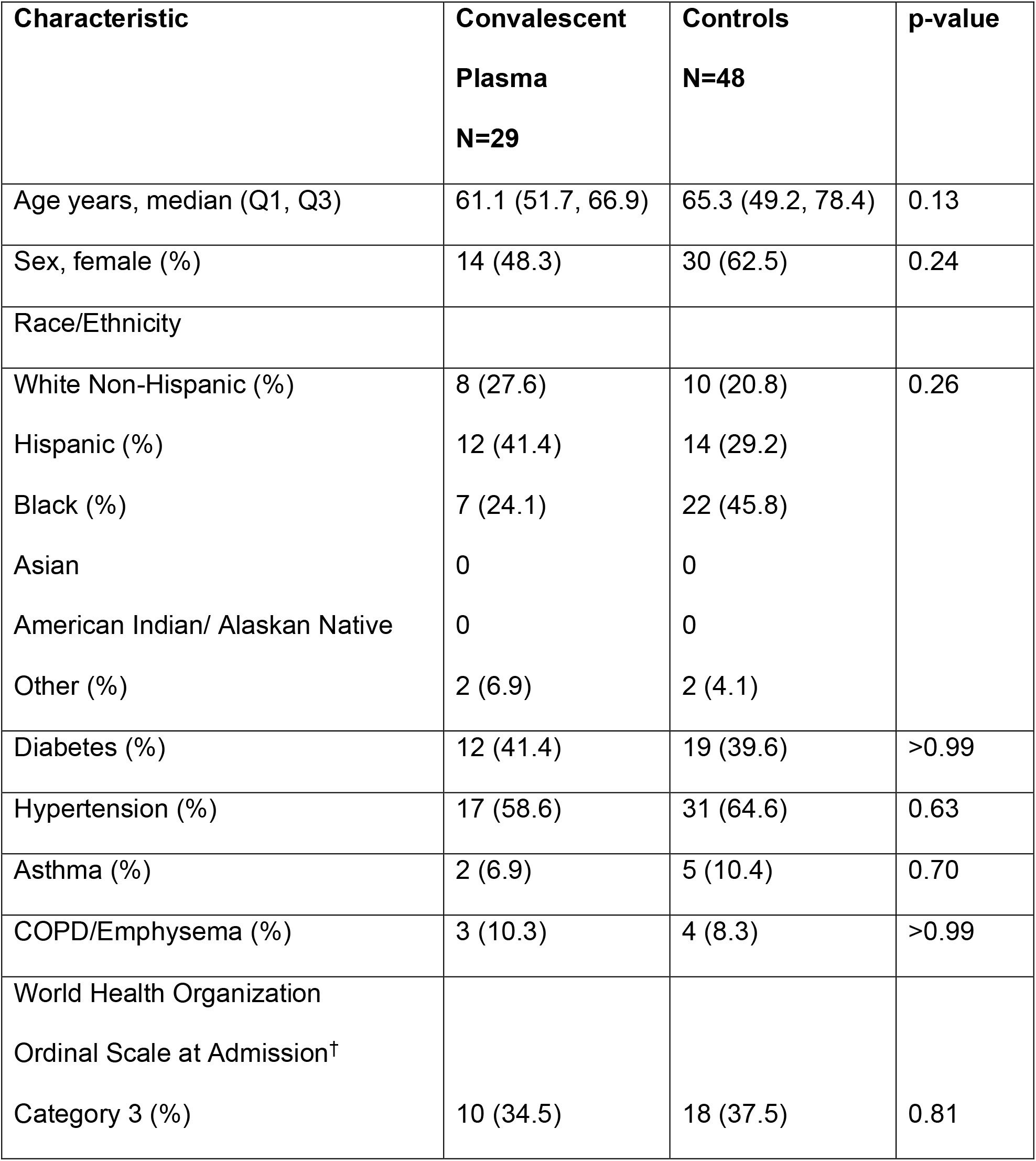
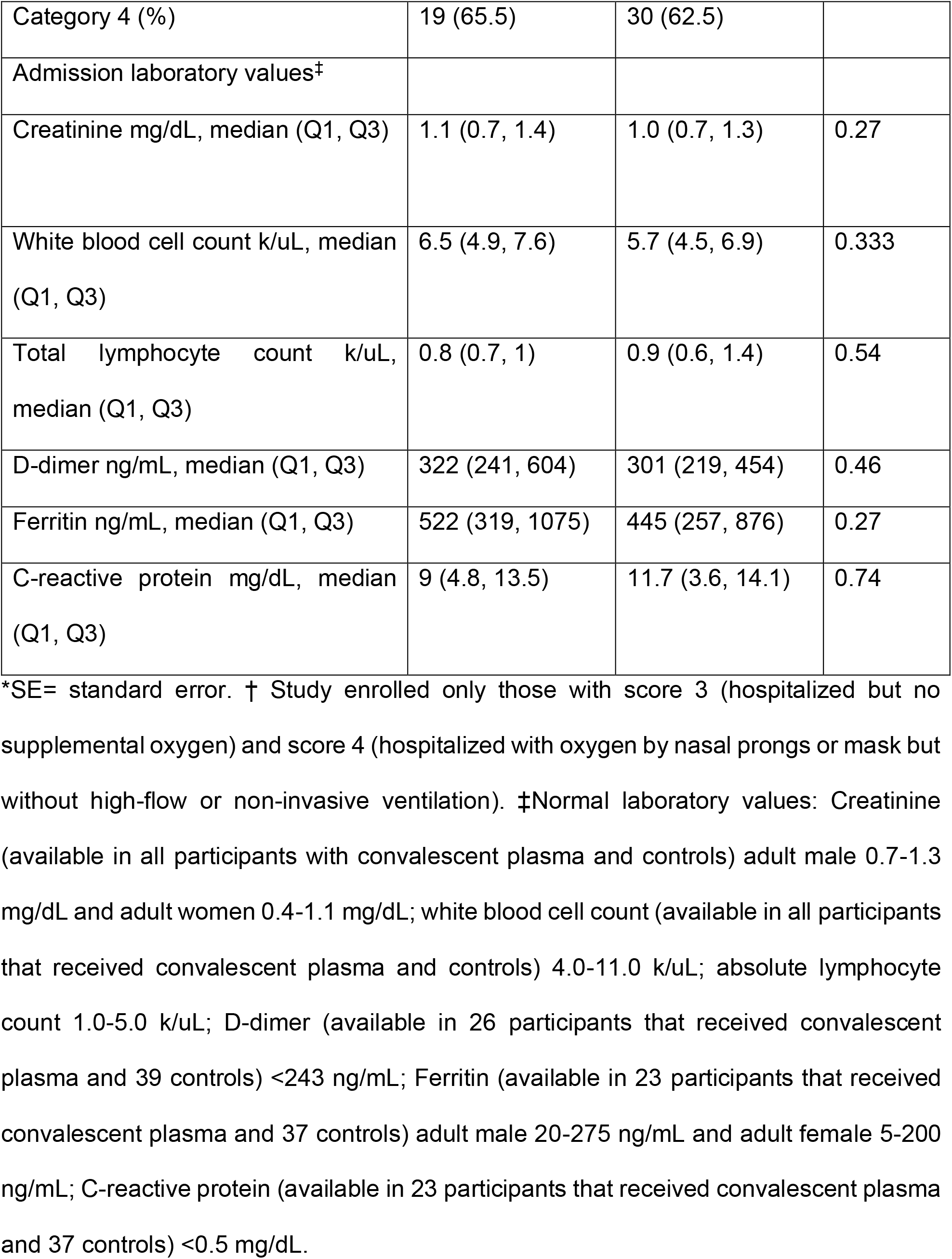
Demographic and Clinical Characteristics Among Participants Receiving Convalescent Plasma and Controls

Of the 29 participants that received CIP, 13 (45%) received their first transfusion within 24 hours of admission, 14 (48%) within 48 hours, and 2 (6%) within 72 hours. One participant (3%) received only one unit of CIP, while the remainder received two units; in 5 (17%) participants, limited supply of available CIP necessitated administration of two units from different donors. All transfused units had detectable IgG to the spike protein, with a considerable range in titer (Table 2, Supplemental Figure 1).

**Table 2.** Immunological profile of the transfused convalescent plasma units.

|  | IgG | IgM | IgA |
| --- | --- | --- | --- |
| Anti-spike µg/ml, median (min-max) | 7.7 (0.1-112.1) | 3.0 (0-106.6) | 2.9 (0-24.7) |
| Anti-RBD µg/ml, median (min-max) | 2.7 (0.1-83.9) | 2.9 (0-27.7) | 2.6 (0-23.5) |
| Anti-nucleocapsid EU/ml, median (min-max) | 0.52 (0.0-8.67) | 1.3 (0-10.0) | 0 (0-2.3) |

\*All units were also screened by the commercial Abbott assay which measures IgG to nucleocapsid but is reported as signal to cut-off values. Per manufacturer recommendation, signal to cut-off values of 1.4 or greater were used to screen. 39 of 42 paired units screened had signal to cut-off values of 1.4 or greater, with range of transfused units from 2.29-9.33.

Overall, CIP transfusion was well-tolerated, similar to prior reports (15). There were 24 adverse events among 11 participants (Supplemental Table 1). Of the serious adverse events, 7 (29%) were grade 3 or above, but none were categorized as related to the transfusion. There were no events of transfusion-related acute lung injury (TRALI) or transfusion-associated circulatory overload (TACO).

The clinical course of each participant is visually represented in the swimmer’s plots depicted in Figure 1. Kaplan-Meier curves for survival and ICU-free survival are represented in Figure 2. We observed a non-statistically significant reduction in the primary endpoint of ICU transfer, with 14% of transfused patients ultimately requiring ICU transfer compared to 27.1% for controls (Fisher’s exact P-value = 0.258). The second primary endpoint of 28-day mortality was similarly non-significantly reduced in the study group at 6.9% compared to 10.4% in controls. A univariate Cox regression analysis for time-to-death (Table 3) showed a hazard ratio (HR) of 0.640 (95% CI 0.124-3.298). Due to the low event rate, no multivariate analysis was performed. Of the other variables tested, only age was significantly associated with mortality, with a HR of 1.103 (95% CI 1.047-1.162). With regard to ICU transfer rate, the univariate time-to-event analysis revealed a HR of 0.506 (95% CI 0.165-1.554) (Table 4). Univariate analyses again showed a weak but statistically significant association of ICU transfer with age, HR 1.03 (95% CI 1.000-1.061). On multivariate analysis (Table 5), the HR for CIP remained similarly non-significantly reduced at 0.470 (95% CI 0.138-1.597). In this analysis, dexamethasone use was found to be significantly associated with risk of ICU transfer (HR 3.217, 95% CI 1.007-10.284), and this is likely reflective of the more severely ill patients receiving dexamethasone, consistent with local standard practice following the results of the RECOVERY trial (16).

**Figure 1.**
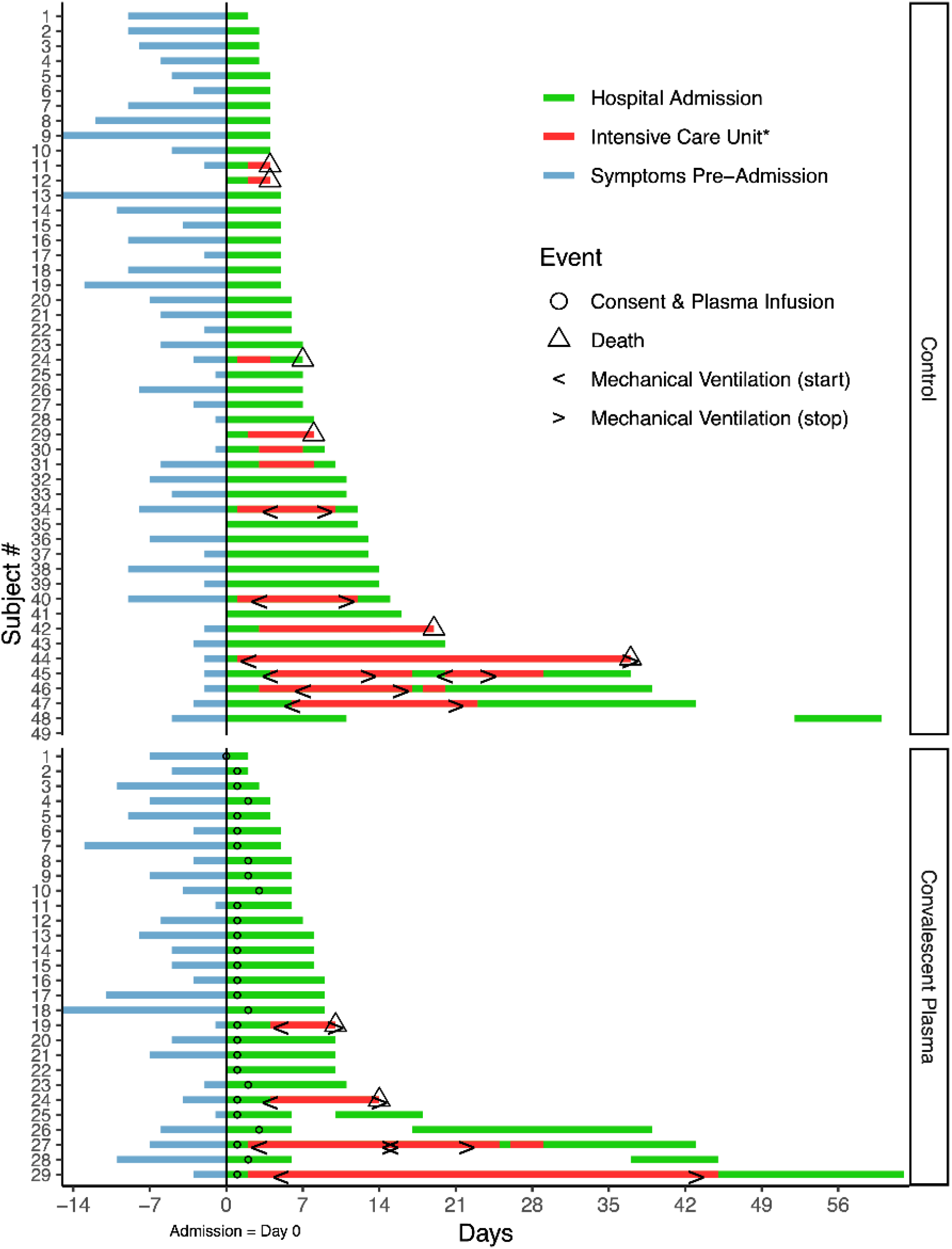
Swimmers plots depicting clinical timelines of CIP transfused participants and controls. Baseline (day 0) was the day of index hospital admission. The blue line represents symptomatic days before admission; green lines represent admission to acute hospital care, with intensive care unit stays represented in red. Blank gaps between hospitalizations indicate the patient was discharged then readmitted within the 60 day followup period. Circles show the date of plasma infusion; triangles indicate that the patient died; “< >” bracket time periods where the patient received mechanical ventilation. Participant 29 in the Convalescent Plasma group was discharged on day 60. *Pictured intensive care unit stays were indicated for higher levels of oxygen therapy including high-flow nasal cannula oxygen, mechanical ventilation, and/or extracorporeal membrane oxygenation.

**Table 3.**
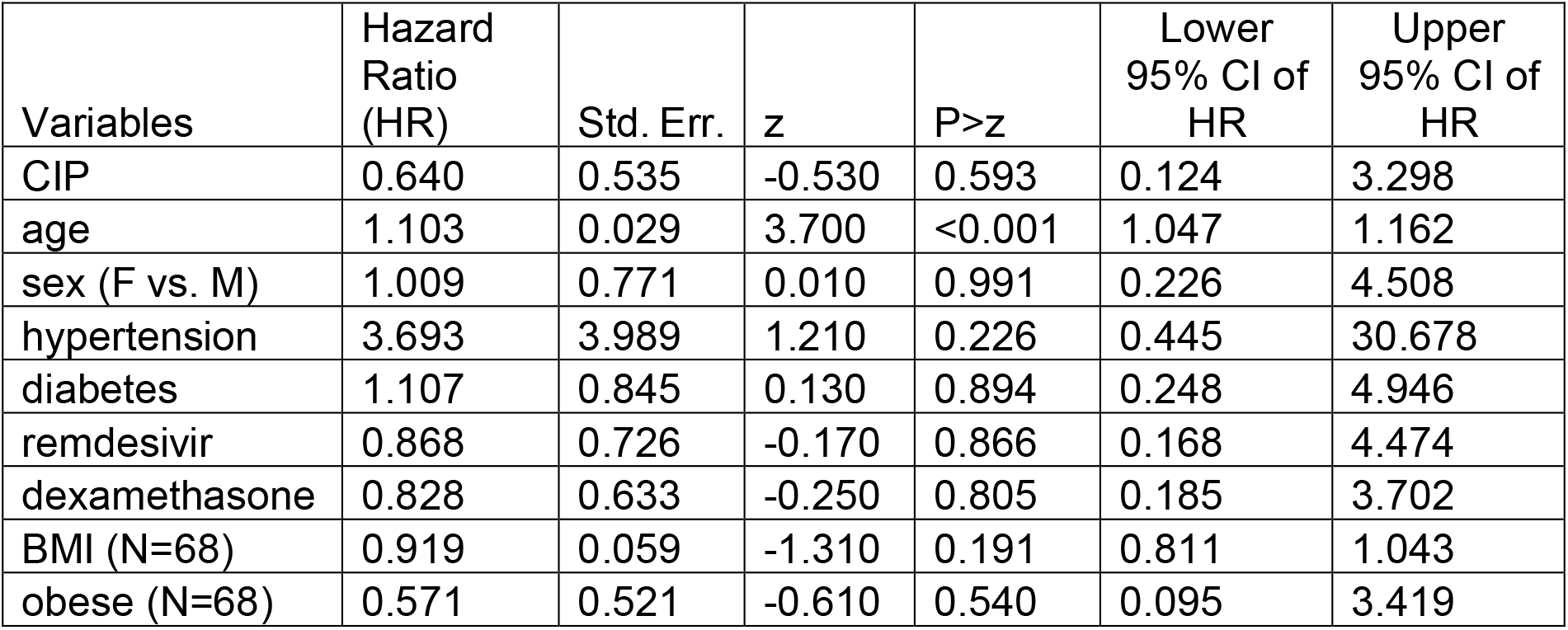
Univariate Cox regression analysis for time to death by 28 days after entering study (N=77)

**Table 4.** Univariate Cox regression analysis for time to ICU (N=77)

| Variables | Hazard Ratio (HR) | Std. Err. | z | P-value | Lower 95% CI of HR | Upper 95% CI of HR |
| --- | --- | --- | --- | --- | --- | --- |
| CIP | 0.506 | 0.290 | -1.190 | 0.234 | 0.165 | 1.554 |
| age | 1.030 | 0.015 | 1.990 | 0.047 | 1.000 | 1.061 |
| sex (F vs. M) | 1.058 | 0.522 | 0.120 | 0.908 | 0.403 | 2.781 |
| hypertension | 1.091 | 0.554 | 0.170 | 0.864 | 0.403 | 2.949 |
| diabetes | 0.946 | 0.466 | -0.110 | 0.911 | 0.360 | 2.487 |
| remdesivir | 1.242 | 0.631 | 0.430 | 0.669 | 0.459 | 3.360 |
| dexamethasone | 2.344 | 1.190 | 1.680 | 0.093 | 0.867 | 6.342 |
| BMI (N=68) | 1.023 | 0.026 | 0.880 | 0.381 | 0.973 | 1.075 |
| obese (N=68) | 1.619 | 1.045 | 0.750 | 0.456 | 0.457 | 5.736 |

**Table 5.** Multivariable Cox regression analysis for time to ICU (N=77)

| Variables | Hazard Ratio (HR) | Std. Err. | z | P-value | Lower 95% CI of HR | Upper 95% CI of HR |
| --- | --- | --- | --- | --- | --- | --- |
| CIPT | 0.470 | 0.293 | -1.210 | 0.226 | 0.138 | 1.597 |
| age | 1.032 | 0.017 | 1.910 | 0.056 | 0.999 | 1.066 |
| sex (F vs. M) | 0.785 | 0.413 | -0.460 | 0.646 | 0.280 | 2.204 |
| hypertension | 0.705 | 0.409 | -0.600 | 0.547 | 0.226 | 2.200 |
| diabetes | 1.270 | 0.709 | 0.430 | 0.668 | 0.425 | 3.791 |
| remdesivir | 0.590 | 0.346 | -0.900 | 0.368 | 0.187 | 1.861 |
| dexamethasone | 3.217 | 1.907 | 1.970 | 0.049 | 1.007 | 10.284 |

**Figure 2.**
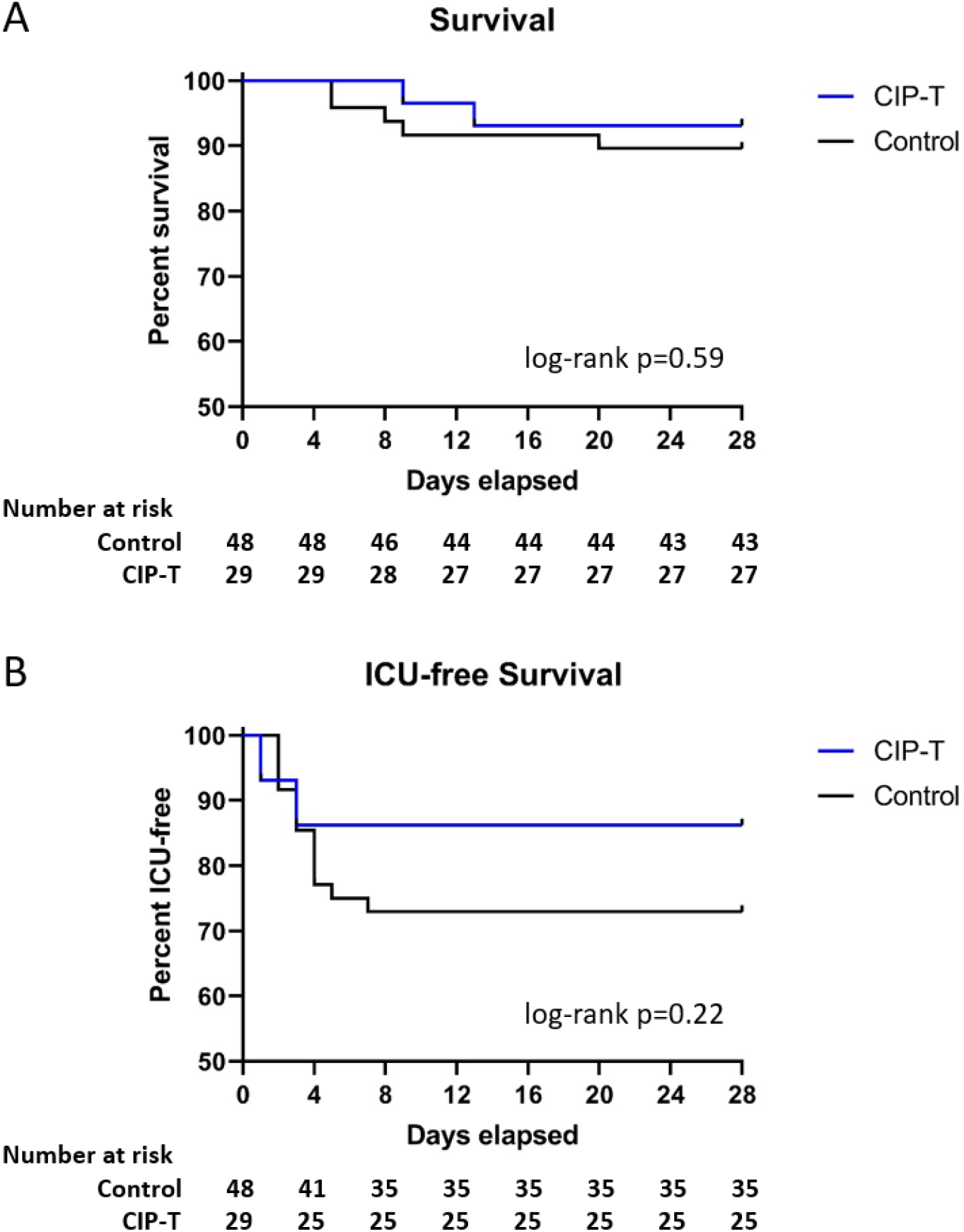
Effect of CIP on progression to critical illness and survival. Kaplan-Meier curves are shown comparing survival (A) and ICU-free survival (B) in CIP transfused patients vs control. Number of patients remaining at risk are listed along the bottom of each panel. Log-rank p values are listed.

Secondary clinical outcomes are summarized in Table 4. There was no significant effect of CIP on any of the clinical secondary endpoints, although most differences between groups showed a similar signal toward benefit of CIP.

### Quantification of Specific Anti-SARS-CoV-2 Antibodies

#### Levels of Specific Antibody in CIP

The levels of specific IgG, IgM, and IgA to spike (S), receptor binding domain (RBD), and nucleocapsid (NC) were measured to determine the levels of each class of specific antibody in the CIP transfused (Table 2, Supplemental Figure 1, panel A). The amounts of IgG anti-NC in the transfused units documented with the Abbott assay (S/CO) vs levels with specific ELISA are available in Supplemental Figure 1, panel B.

#### Specific Antibody Levels in Participants

In order to determine the kinetic pattern of specific SARS-CoV-2 IgG, IgM, and IgA responses to S, RBD, and NC after CIP transfusion, specific levels of IgG, IgM, and IgA were measured at baseline, and on days 7, 14, and 28 after CIP transfusion. All classes of antibodies to the three SARS-CoV-2 target proteins were significantly increased at days 7 and 14 post-transfusion compared to baseline (Figure 3, p<0.01). Anti-NC IgG levels were reduced at day 28, suggesting that the initial rise was likely due in part to the contribution of the transfused units. Supplemental Table 2 summarizes the distribution for all specific antibodies measured over time.

**Figure 3:**
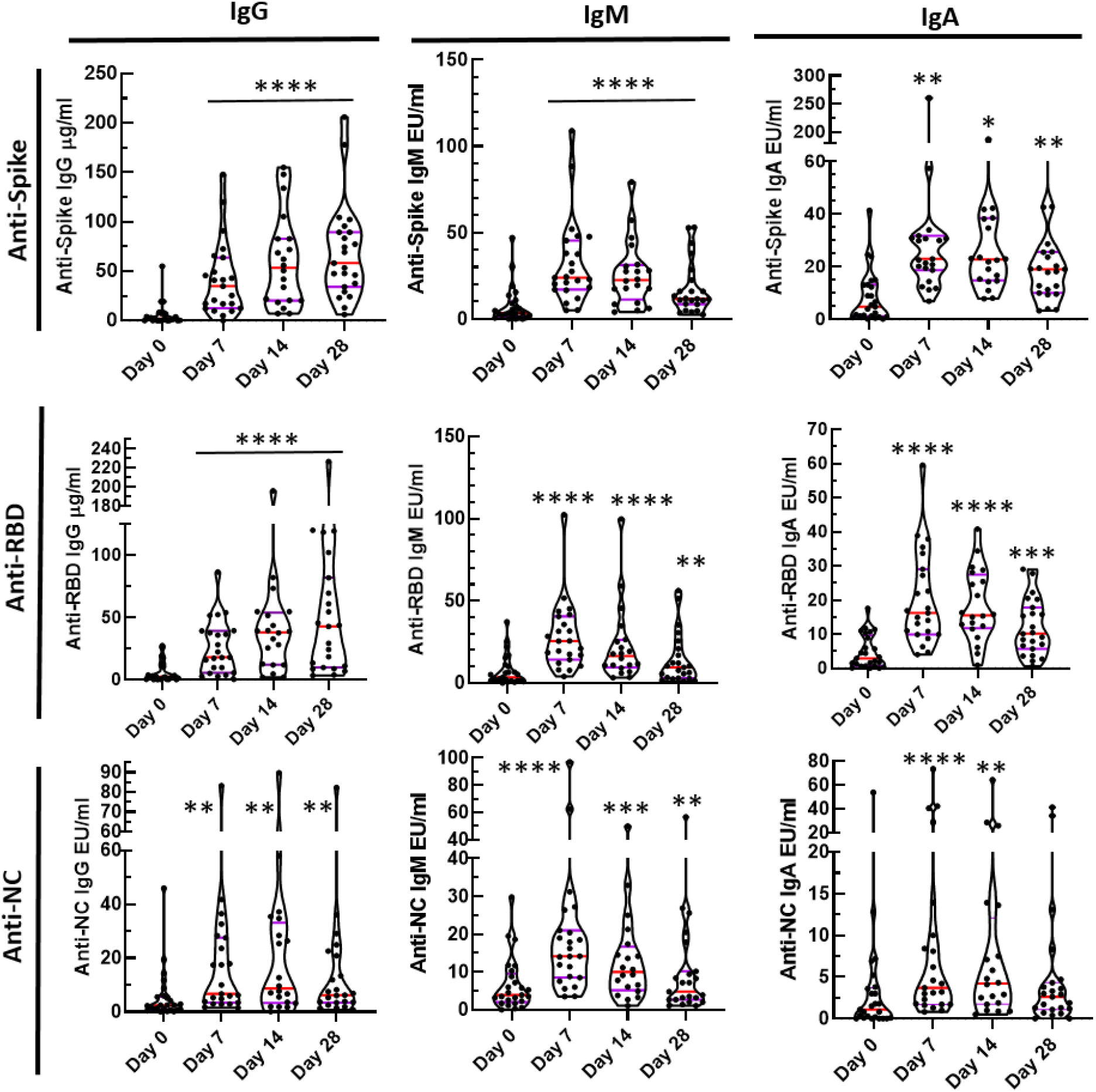
Specific IgG, IgM, and IgA antibodies directed at spike (S), receptor binding domain (RBD), nucleocapsid (NC). Blood was collected on CIP treated participants immediately prior to infusion (day 0), and then 7, 14, and 28 days post-infusion. Levels of specific anti-SARS-CoV-2 antibodies were measured and compared to baseline. Paired Wilcoxon rank sum p values: * <0.02; **< 0.01; *** <0.001; **** < 0.0001. Medians and the 25 and 75 quartiles are indicated on each violin plot.

The median anti-spike IgG, IgM, and IgA levels were significantly increased over baseline at days 7, 14, and 28 (p<0.0001). The median IgM anti-S response was markedly elevated at days 7 and 14 but was decreased by day 28. The median anti-RBD IgG, IgM, IgA levels rose significantly (p<0.0001) with the IgG anti-RBD levels that persisted at day 28 while IgM levels began to decrease by day 14 with significant reduction but clearly detectable levels by day 28. In the case of IgM anti-S, -RBD, and NC, the decrease of IgM may reflect IgG to IgM class switching. Correlation analyses were performed testing the relationship between CIP and post-transfusion circulating specific antibody levels at day 7, but no significant associations were found. Analyses to test the association of CIP specific antibody levels with time to ICU transfer and time to viral PCR negativity were also performed. Only one significant association was found (Supplemental Table 3): a positive one between CIP anti-RBD IgM and time to ICU transfer. This result is interpreted with caution since it was a single ICU case for which the CIP anti-RBD IgM level was markedly elevated.

### Respiratory Tract Viral Clearance

In order to better understand the kinetics of respiratory tract viral clearance, nasopharyngeal swabs were collected at baseline and days 4, 7, 14, and 21 post-transfusion. SARS-CoV-2 PCR cycle thresholds over time for transfused patients are represented in Figure 4, with those patients transferring to the ICU denoted in red. Kaplan-Meier survival curves showed a statistically non-significant reduction in time to first negative PCR with CIP compared with 19 controls with >1 PCR test following admission (mean 20.4 vs. 24.8 days, log-rank test p=0.22, Supplemental Figure 2). There was no association between CIP anti-SARS-CoV-2 specific antibody levels and time-to-PCR negativity (Supplemental Table 4).

**Figure 4.**
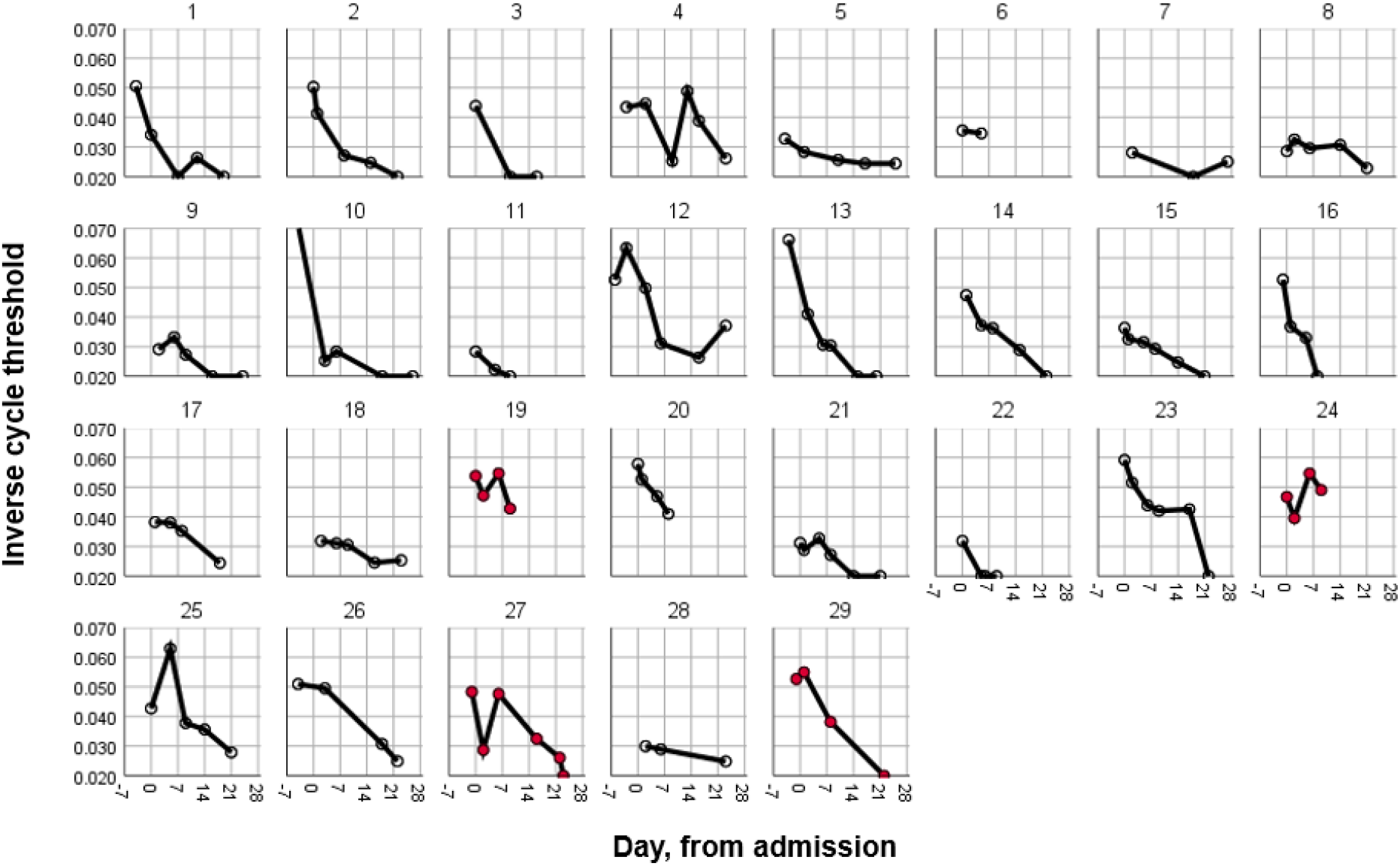
Respiratory Tract Viral Clearance. Serial respiratory tract swabs were collected at baseline (day 0) and then 4, 7, 14, and 21 days post CIP transfusion. The inverse cycle thresholds for SARS-CoV-2 RNA are graphed. Participants who were ultimately transferred to the ICU are depicted in red.

Understanding the clinical factors which influence viral clearance may also provide insight into disease pathogenesis and mechanisms of recovery, regardless of the effect of CIP. Therefore, we used these data to test for associations between clinical variables and time-to-PCR negativity (Supplemental Tables 5 and 6). Of note, hypertension and Angiotensin Converting Enzyme Inhibitor (ACEI) use were the only variables significantly associated with a significantly longer time to negativity (in univariate and multivariate analyses).

## DISCUSSION

In this study, we report the effect of CIP on the progression to critical illness in people with severe COVID-19. To our knowledge this is the first study of its kind to target severe, non-critically ill patients, and to provide a detailed analysis of the kinetics of anti-SARS-CoV-2 specific antibody levels and viral shedding following CIP infusion. This study was implemented at the beginning of the pandemic when estimates for effect size were limited by early available epidemiologic data suggesting that roughly 50% of admitted patients necessitated ICU transfer. Ultimately, this phase 2 single-arm study was underpowered for the primary endpoint of ICU transfer, as local baseline rates of ICU transfer proved to be lower than initially predicted for the population admitted. Kaplan-Meier curves trend in the direction of CIP infusions benefiting the participants but were limited by the sample size included in this study. Based on our observed control ICU transfer rate of 27%, a subsequent study in a similar population would require approximately 166 patients per group (N=332 total) to have 80% power to detect a 50% reduction in ICU transfer.

Anti-SARS-CoV-2 specific IgG, IgM, and IgA antibodies rose to S, RBD, and NC from baseline to post-transfusion, with highly significant differences between baseline and days 7 and 14 post-transfusion. There was little change in IgG anti-S and anti-RBD between day 7 and day 28 post-infusion in most cases, suggesting that transfused CIP antibodies persisted while endogenous IgG and IgA were being made. In contrast, the reductions in specific IgM levels to the various proteins suggests that IgG to IgM switching was occurring leading to rapid decreases in IgM anti-S and anti-RBD antibody levels by day 28. These kinetics differ significantly from that reported elsewhere for non-transfused patients in whom peak IgG levels without CIP occur at 3-4 weeks post-infection (17) and suggest that CIP infusions significantly impacted early post-transfusion circulating anti-SARS-CoV-2 specific antibody levels. In support of this, in an early small case series of 5 critically ill patients, Shen et al (18) showed that RBD specific IgG and IgM and neutralizing antibody titers rose between day 0 and 3 after CIP infusions and remained at the same levels through day 7. Similarly, in a recent study of CIP in older adults with severe COVID-19, Libster *et al*. reported a significant difference in circulating anti-SARS-CoV-2 spike IgG levels between placebo and CIP infused patients at 24 hours post-infusion (19). The quantitation of specific levels of monoclonal antibodies in patients who receive monoclonal antibodies that could be tracked with an anti-idiotypic antibody would provide further insight into persistence of passive antibody infusions and the amounts of endogenously produced specific antibodies. Additionally, future studies examining antibody secretion *in vitro* from circulating B cells isolated from CIP transfused participants could also provide insights into the relative contributions of exogenous and endogenous antibodies.

We did not find a relationship between CIP and post-transfusion circulating specific antibody levels at day 7, likely reflecting variability in CIP antibody levels and function, variable decay of the infused antibodies, and variable rates of endogenous production. We also did not observe a statistically significant effect of CIP on viral clearance, which again may be reflective of variable contributions of host antiviral immunity (be that cellular or humoral) and CIP. Unrelated to CIP transfusion, patients with hypertension or on an ACE inhibitor had significantly longer time to PCR negativity, which is in keeping with other reports in the literature (20). One proposed explanation for this could be up-regulation of ACE2 receptors in these patients, the cellular target of SARS-CoV-2.

Passive antibody therapy is considered most effective when used for prophylaxis of infectious diseases, and delay of administration in active disease is associated with waning efficacy over the course of infection. For example, antibody therapy for pneumococcal pneumonia was most effective when given shortly after the onset of symptoms and was of no benefit beyond the third day of disease (21) and similar effects were seen in CIP trials for SARS-CoV-1 and H1N1 influenza (4). Available evidence suggests this may also be the case with COVID-19. For example, the retrospective study by Liu et al. found that the benefit of CIP was limited to those patients with symptom onset less than or equal to 7 days prior to admission and those not intubated, suggesting that patients treated earlier in the course of disease before the onset of critical illness may derive the most benefit (22). Fittingly, in the largest data set available from the EAP (23), both early administration and higher CIP titers were associated with improved outcomes. Lastly, in addition to the timing of treatment, patient-specific factors may help determine who is more likely to benefit. A recent study focusing on adults with mild disease who were greater than 75 years of age or 65 with comorbid conditions found that CIP was protective of progression to severe disease in this group (19). An important socioeconomic feature of our study is that it included a majority of participants that identified as Black race or Hispanic ethnicity, groups which are historically underrepresented in clinical trials and have been disproportionately affected by the COVID-19 pandemic. Similar inclusion of representative groups in future studies will be critical to understanding the generalizability of the effects of CIP on COVID-19 disease progression (24, 25).

As this pandemic continues to evolve, so too must the treatments. The emergence of new strains of SARS-CoV-2 raises the specter of potential resistance to developing therapies, including specific monoclonal antibodies. In this setting, evidence suggests that polyclonal CIP with its array of antibodies targeting different viral proteins remains an available and importantly adapting therapeutic alternative.

## Data Availability

De-identified source data can be provided upon request once appropriate permissions are obtained as per institutional review board regulations.

## ACKNOWLEDGMENTS

The authors would like to acknowledge Bruce Sachals, MD, PhD, Chief Medical Officer and Vice President of the New York Blood Center Enterprises, and Professor of Pathology and Laboratory Medicine at Weill Cornell Medical College for supplying CIP. The authors acknowledge the technical efforts of Dana Schalk, Amy Schienschang, Amanda Polend, Abdalla Elhakiem, Sarah Whitaker, and Michael LaBrie for running the specific antibody titers.

