## supplemental material for "Convalescent plasma for preventing critical illness in COVID-19: A phase 2 trial and immune profile"

### SUPPLEMENTAL METHODS

#### Convalescent Plasma

CIP was provided by the New York Blood Center. Units were prescreened by semi-quantitative IgG to the nucleocapsid protein using the Abbott SARS-CoV-2 chemiluminescence enzyme immunoassay (Architect i2000 chemiluminescent microparticle assay; Abbott; Abbott Park, IL). Per manufacturer, a signal to cut-off (S/CO) value of 1.4 or greater was used to define presence of antibody; the range of S/CO values among CIP units transfused was 2.3 to 9.3. Notably, the use of the Abbott assay, and its S/CO cutoff, precedes the FDA's recommendation within the EUA that CIP be screened for "high titer" anti-spike levels using the Ortho VITROS SARS-CoV-2 IgG assay (Ortho S/CO ≥ 12), but titers to these targets have been shown to correlate (1, 2). CIP was administered as two paired units from the same donor, when available, (~220 mL each) over 1-2 days. Prior to subject transfusion, aliquots of CIP units were taken and stored at 4°C for testing ELISA specific IgG, IgM, and IgA to the spike (S), receptor binding domain (RBD) nucleocapsid (NC) proteins. Spike (ProSci Inc, San Diego, CA), Nucleocapside (ProSci), and RBD (Rayriotech, Reachtree Corner, CA) antigens, and secondary developing antibodies for IgG, IgM, and IgA (Jackson ImmunoResearch Laboratories, Inc, West Grove, PA) were tested for specificity and pre-titrated for optimum dose and time using well-described methods for antibody-specific ELISAs. CIP and sample from different time points were processed, stored, and tested in the same manner with reference controls used to normalize the values between runs and samples.

**Laboratory Measurements and Clinical Follow-up**

At enrollment, demographics, comorbidities and symptoms were collected along with the date of symptom onset. Vital signs were recorded along with oxygen saturation, oxygen requirement, and laboratory values including neutrophil count, lymphocyte count, C-reactive protein (CRP), fibrinogen, and d-dimer. Results of chest imaging were recorded but not specifically performed for the trial. For contemporaneous controls, similar data were extracted from the medical record.

Safety assessments were performed on the day of transfusion (day 0), and on days 1, 2, 3, 4, 7, 14, 21, and 28 and 60. Adverse events were labeled and scored using the Common Terminology Criteria for Adverse Events version 5.0 using the severity scale: 1, Mild: transient or mild discomfort (<48 hours); no medical intervention/therapy required); 2, Moderate: some worsening of symptoms but no or minimal medical intervention/therapy required; 3, Severe: escalation of medical intervention/therapy required; 4, Life-threatening: marked escalation of medical intervention/therapy required; 5, Death (3). The primary endpoint of ICU transfer was considered to have been met for any participant that was physically moved to an ICU-level monitored bed and received critical care as defined by consistent supplemental oxygen of >6 liters per minute at rest, high-flow nasal cannula, non-invasive positive pressure, mechanical ventilation, or vasopressors. In some instances, the participant qualified for ICU transfer based on the above criteria but based on his/her goals level of care was not escalated. These cases were adjudicated as having progressed to critical illness. Serum antibody titers to SARS-CoV-2 total IgG, IgM, and IgA to the spike, nucleocapsid and RBD proteins were completed on days 0, 7, 14, and 28. SARS-CoV-2 PCR from nasopharyngeal swabs were performed at days 0, 4, 7, 14 and 21 for CIP recipients; for contemporaneous controls, all available SARS-CoV-2 PCR results collected for clinical purposes were recorded. PCR assays were performed with either the Xpert Xpress SARS-CoV-2 (Cepheid, Sunnyvale CA, USA), Alinity mSARS-CoV-2 (Abbott Molecular, Des Plaines, IL), or Abbott M2000 RealTime SARS-CoV-2 assay (Abbott, Chicago, IL) using reagents and protocols according to the Emergency Use Authorization approved assays for SARS CoV-2 detection. When assessing cycle thresholds (CT), results performed on the M2000 platform were adjusted by ten cycles to ensure comparability between platforms. When needed, the lower of the two measurements (i.e. CT to detect the N2 gene targets or CT to detect the E gene targets) was used.

**Statistical Analyses**

Descriptive statistics for cohort demographic variables were calculated as reported in the results. To compare patients who received CIP versus controls, Wilcoxon rank-sum test was used for continuous variables and Fisher's exact test was used for categorical variables. For time-to-event data, Kaplan-Meier curves were generated and compared with log-rank test. For the primary endpoint of mortality, due to the low number of events a univariate Cox regression was performed for CIP as well as other relevant variables of interest which included age, sex, hypertension, diabetes, remdesivir, dexamethasone, BMI, and obesity (defined as BMI $\geq$30). Similarly, univariate Cox regression analyses were performed for time-to-ICU data. Multivariable Cox regression analysis was also performed to explore the association between CIP and time-to-ICU adjusting for above mentioned relevant variables. Similar analyses were carried out for time-to-negative PCR. To compare the change of specific antibody measurements following CIP transfusion from baseline, paired non-parametric method Wilcoxon signed rank test was applied for each follow-up date, respectively. Pearson correlation coefficients were calculated and tested for the correlation between CIP and post-transfusion circulating antibody levels.

### SUPPLEMENTAL TABLES

**Supplemental Table 1**: Participants reporting at least 1 adverse event

| Adverse Event | Convalescent Plasma Recipients (N=29) |
|---|---|
| Any event, n (%)* | 11 (38) |
| Related adverse event | 4 (14) |
| Severe (≥ grade 3) | 7 (24) |
| Life-threatening (grade 4) | 3 (10) |
| Any serious adverse event | 4 (14) |
| Related serious adverse event** | 0 |
| Severe (≥ grade 3) | 4 (14) |
| Life-threatening (grade 4) | 3 (10) |
| Death | 2 (7) |

* Unique adverse events included pulmonary edema (n=6 occurrences in 5 participants), acute respiratory failure (n=4), fever (n=3), hypoxia (n=1) elevated aspartate aminotransferase (n=1), elevated alanine aminotransferase (n=1) atrial fibrillation with rapid ventricular response (n=1), elevated creatinine (n=1), epistaxis (n=1), hypertensive urgency (n=1), hypotension (n=1), sepsis (n=1), worsening sepsis (n=1), syncope (n=1).

**A related event was categorized as definitively, probably or possibly related.

**Supplemental Table 2**. Comparison of specific antibodies in CIP recipients over time.

| | | Day 0 | Day 7 | | Day 14 | | Day 28 | |
|---|---|---|---|---|---|---|---|---|
| | | | | p value | | p value | | p value |
| **Spike** | **IgG** | **1.9** (0.065-6.6) | **35.0** (13.0-64.0) | <0.0001 | **53.0** (20.0-83.0) | <0.0001 | **58.0** (34.0-90.0) | <0.0001 |
| | **IgM** | **3.4** (1.45-10.0) | **23.9** (17.10-45.4) | <0.0001 | **22.55** (11.33-31.33) | <0.0001 | **11.70** (8.5-22.50) | 0.0002 |
| | **IgA** | **4.8** (1.15-13.15) | **23.0** (18.7-31.8) | <0.0001 | **22.75** (14.78-38.5) | <0.0001 | **19.0** (10.0-25.7) | <0.0001 |
| **RBD** | **IgG** | **2.26** (0.28-10.8) | **17.9** (5.54-39.13) | <0.0001 | **38.1** (12.02-53.87) | <0.0001 | **42.8** (9.68-81.82) | <0.0001 |
| | **IgM** | **3.4** (1.4-10.6) | **25.6** (14.3-40.6) | <0.0001 | **16.4** (9.6-26.25) | <0.0001 | **9.6** (3.1-20.9) | 0.0037 |
| | **IgA** | **2.9** (0.7-9.6) | **16.2** (9.9-29.0) | <0.0001 | **15.5** (11.73-27.43) | <0.0001 | **10.1** (5.7-17.8) | 0.0002 |
| **NC** | **IgG** | **2.4** (0.9-5.7) | **6.7** (3.4-27.50) | 0.0022 | **8.7** (3.3-33.13) | 0.0067 | **6.2** (3.4-20.90) | 0.0033 |
| | **IgM** | **3.9** (2.0-9.55) | **14.2** (8.5-21.0) | <0.0001 | **10.0** (5.2-16.68) | 0.0005 | **4.8** (2.7-10.2) | 0.0079 |
| | **IgA** | **1.1** (0.1-3.7) | **3.7** (1.7-10.0) | <0.0001 | **4.2** (1.75-12.08) | 0.0024 | **2.6** (1.1-4.3) | 0.6320 |

Wilcoxon signed-rank test was used to compare Day 0 levels with day 7, 14, and 28 levels of each specific antibody. Median with the 25$^{th}$ and 75$^{th}$ percentiles are listed.

**Supplemental Table 3**. Univariate Cox regression analysis examining the correlation between CIP antibody levels and time to ICU transfer (N=25).

| Variable | Hazard Ratio (HR) | Std.Err. | z | P-value | Lower bound 95% CI of HR | Upper bound 95% CI of HR |
|---|---|---|---|---|---|---|
| IgG Spike CIP | 1.014 | 0.01521 | 0.911 | 0.362 | 0.984 | 1.044 |
| IgG RBD CIP | 1.008 | 0.02256 | 0.371 | 0.711 | 0.965 | 1.054 |
| IgG NC CIP | 0.949 | 0.261 | -0.192 | 0.848 | 0.553 | 1.626 |
| IgM Spike CIP | 1.017 | 0.01451 | 1.206 | 0.228 | 0.989 | 1.046 |
| IgM RBD_CIP | 1.144 | 0.06807 | 2.254 | 0.024* | 1.018 | 1.285 |
| IgM NC CIP | 1.190 | 0.1885 | 1.098 | 0.272 | 0.872 | 1.623 |
| IgA Spike CIP | 1.042 | 0.04789 | 0.901 | 0.367 | 0.953 | 1.140 |
| IgA RBD CIP | 1.058 | 0.08371 | 0.711 | 0.477 | 0.906 | 1.235 |
| IgA NC_CIP | 1.920 | 1.016 | 1.234 | 0.217 | 0.681 | 5.415 |

**Supplemental Table 4:** Univariate Cox regression analysis examining the correlation between CIP antibody levels and time to PCR negativity (N=25).

| Variable | Hazard Ratio (HR) | Std.Err. | z | P-value | Lower bound 95% CI of HR | Upper bound 95% CI of HR |
|---|---|---|---|---|---|---|
| IgG Spike CIP | 1.014 | 0.009 | 1.554 | 0.120 | 0.996 | 1.033 |
| IgG RBD CIP | 1.006 | 0.015 | 0.423 | 0.672 | 0.978 | 1.036 |
| IgG NC CIP | 0.924 | 0.116 | -0.635 | 0.526 | 0.723 | 1.181 |
| IgM Spike CIP | 0.970 | 0.038 | -0.785 | 0.432 | 0.899 | 1.047 |
| IgM RBD CIP | 0.993 | 0.044 | -0.148 | 0.882 | 0.910 | 1.084 |
| IgM NC CIP | 0.950 | 0.091 | -0.533 | 0.594 | 0.788 | 1.146 |
| IgA Spike CIP | 0.964 | 0.033 | -1.075 | 0.283 | 0.901 | 1.031 |
| IgA RBD CIP | 1.056 | 0.043 | 1.343 | 0.179 | 0.975 | 1.144 |
| IgA NC_CIP | 0.701 | 0.244 | -1.022 | 0.307 | 0.355 | 1.385 |

**Supplemental Table 5.** Univariate Cox regression analysis for time to negative PCR result (N=48)

| Variables | Hazard Ratio | Std. Err. | z | P>z | Lower 95% CI | Upper 95% CI |
|---|---|---|---|---|---|---|
| CIPT | 1.672 | 0.710 | 1.210 | 0.226 | 0.727 | 3.843 |
| age | 0.988 | 0.014 | -0.810 | 0.419 | 0.961 | 1.017 |
| sex (F vs. M) | 1.447 | 0.573 | 0.930 | 0.351 | 0.666 | 3.143 |
| hypertension | 0.274 | 0.114 | -3.120 | 0.002 | 0.122 | 0.619 |
| diabetes | 0.944 | 0.391 | -0.140 | 0.889 | 0.419 | 2.126 |
| BMI | 0.994 | 0.018 | -0.360 | 0.718 | 0.959 | 1.029 |
| obese | 1.925 | 1.065 | 1.180 | 0.237 | 0.651 | 5.694 |
| remdesivir | 0.650 | 0.288 | -0.970 | 0.330 | 0.273 | 1.548 |
| dexamethasone | 0.569 | 0.222 | -1.450 | 0.148 | 0.265 | 1.222 |
| ACE inhibitor use | 0.298 | 0.141 | -2.560 | 0.011 | 0.118 | 0.753 |

**Supplemental Table 6.** Multivariable Cox regression analysis for time to negative PCR result (N=45)

| Variables | Hazard Ratio | Std. Err. | z | P>z | Lower 95% CI | Upper 95% CI |
|---|---|---|---|---|---|---|
| CIPT | 0.597 | 0.309 | -0.990 | 0.320 | 0.216 | 1.649 |
| age | 0.991 | 0.018 | -0.480 | 0.632 | 0.956 | 1.028 |
| gender | 1.247 | 0.548 | 0.500 | 0.616 | 0.527 | 2.949 |
| hypertension | 0.244 | 0.139 | -2.480 | 0.013 | 0.080 | 0.743 |
| diabetes | 2.910 | 1.844 | 1.690 | 0.092 | 0.841 | 10.073 |
| ACE inhibitor use | 0.295 | 0.174 | -2.070 | 0.038 | 0.093 | 0.936 |

### SUPPLEMENTAL FIGURE LEGENDS

**Supplemental Figure 1**: Levels of Specific antibodies in CIP. A) Levels (ELISA Units= EU) in UVA assays for IgG, IgM, and IgA anti-spike, receptor binding domain (RBD) , and nucleocapsid.(NC); B) Levels of IgG anti-NC in Abbott assay with signal/cut-off (S/CO) values and UVA ELISA (μg/ml). Medians and $25^{th}$ and $75^{th}$ percentiles are shown.

**Supplemental Figure 2**. Respiratory tract viral clearance. Serial respiratory tract swabs were obtained on CIP participants at days 0, 4, 7, 14, and 21 post-transfusion. Time to first negative PCR was compared to controls with greater than 1 PCR performed following admission.

**Supplemental Figure 1.**

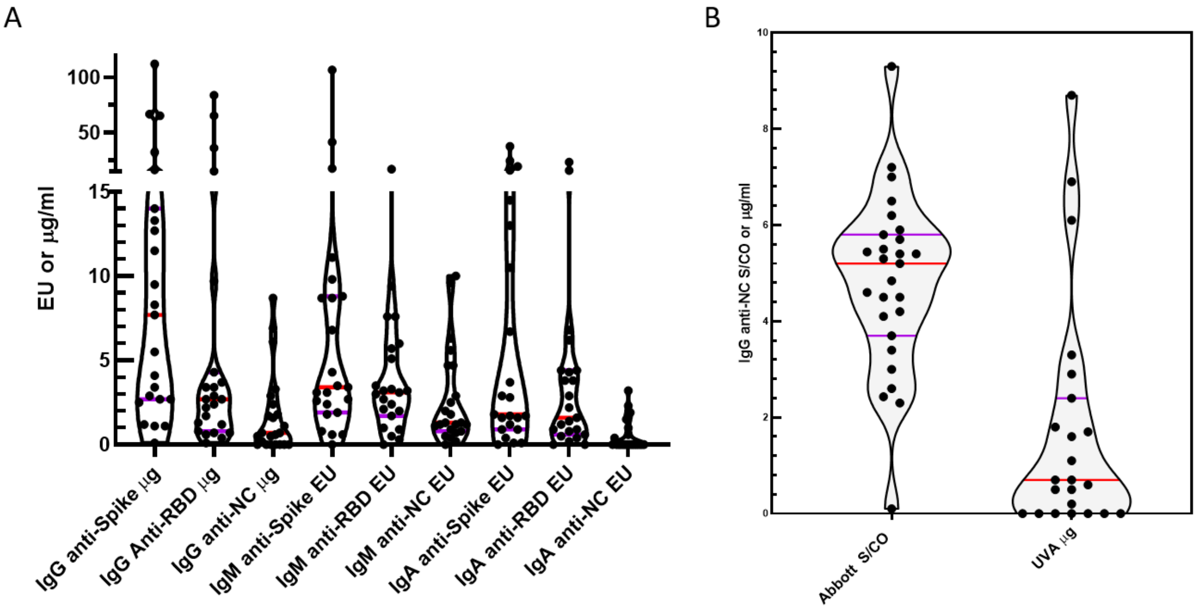

**Supplemental Figure 2.**

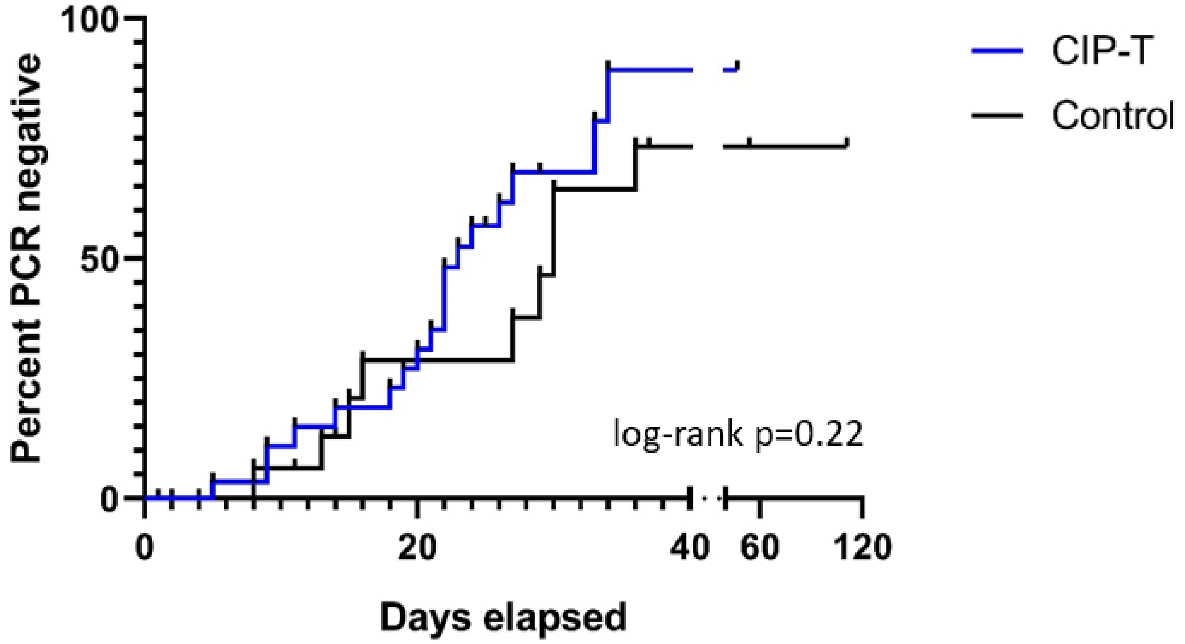
